# The Wearipedia Project: a free and open-source resource for understanding and using wearables in decentralized clinical trials

**DOI:** 10.1101/2025.05.12.25327465

**Authors:** Alexander Johansen, Kyu Hur, Jack Hung, Rodrigo Castellon, Tristan Peng, Stephanie Ren, Renee White, Chanyeong Park, Allison Lau, Saarth Shah, Hee Jung Choi, William Wang, Pann Sripitak, Mohamed Elhusinni, Michael Snyder

## Abstract

**Background:** Finding the optimal wearable biomedical sensor (ref. wearable) for a clinical research study can be challenging. Many wearables are consumer electronics and are not designed for clinical research and their clinical variables vary widely. We aimed to build a resource for clinical researchers to select the best device for their research study, and programming tools to facilitate wearable research.

**Methods:** For each wearable entry, we document the following— <u>Open-source coding tools</u>: we built data extraction, simulation, statistical testing, and educational materials; <u>Clinical trial usage</u>: trials using the device including ChatGPT-generated summaries; <u>Privacy evaluation</u>: low data risk, HIPAA compliance, de-identification, third-party data sharing, and third-party data sharing transparency; <u>Security evaluation</u>: wearable connectivity and API access protocols.

**Findings:** The Wearipedia database consists of 19 wearables + 5 Apps; 7 smart watches, 4 fitness trackers, 2 chest straps, 2 CGM devices, 1 smart ring, 1 arm strap, 1 under the bed sleep tracker, 1 smart scale, 2 apps for diet tracking, 1 app for questionnaires, and 2 apps for data storage. For <u>public coding tools</u>, there where 891 pages of educational material across 22 wearables and apps. We support data extraction from 13 official APIs and 3 unofficial APIs under the Wearipedia pypi package. For <u>clinical usage</u>, there where 63 (± 99) clinical trials per device. For <u>security and privacy</u>, a total of 87 citations and an average of 3.48 citations are referenced, mostly consisting of privacy policies, terms-of-service agreements, and wearable manuals. The Wearipedia database is conveniently accessible through a website at https://wearipedia.com.

**Interpretations:** Wearables can accurately predict important physiological parameters, glucose, and sleep. However, access to high resolution data can be restrictive, characterizing data accuracy is difficult, and wearable data is often not protected from third party reselling, including government requests.

**Funding:** This work was made possible by the support of the BV and Anu Jagadeesh Family Foundation.

## Introduction

Wearable biomedical sensors (i.e. wearables) are a group of electronic devices that can provide continuous, inexpensive, and real-time physiological information using either noninvasive or minimally invasive measurements^1^. Although not generally FDA approved for most applications, individuals, clinicians, and clinical researchers can use wearable sensors to track patient health data and conduct decentralized clinical studies and trials^2,3^. In decentralized clinical studies, the clinical researcher might not have direct contact with the test subject and requires remote collection of health parameters. Wearables provide decentralized clinical studies with a tool that is inexpensive (as low as US$80 per subject), allows 24/7 monitoring, only requires partial internet connectivity, and are designed to be minimally intrusive to the users daily tasks^4,5^. Recent efforts in clinical studies have shown promising results using wearables to predict features such as COVID19 before symptom onset^6,7^, menstrual cycle and pregnancy^8–10^, and abnormal heart function^11^. These advances come through developments in modern sensor technology and signal processing that takes raw sensor output and produces meaningful high-level features through mathematical modeling that researchers can access through wearable manufacture application interfaces (APIs) (ex. Oura, Fitbit) or software development kits (SDKs) (e.g. Health kit). Figure 1 provides a screenshot of a device in the Wearipedia database, namely the Oura Ring gen 3. The website contains detailed information on device capabilities and use, educational material, API tools, and concerns about privacy and security. The Wearipedia website covers the Oura Ring gen 3 and many other popular devices.

**Figure 1:**
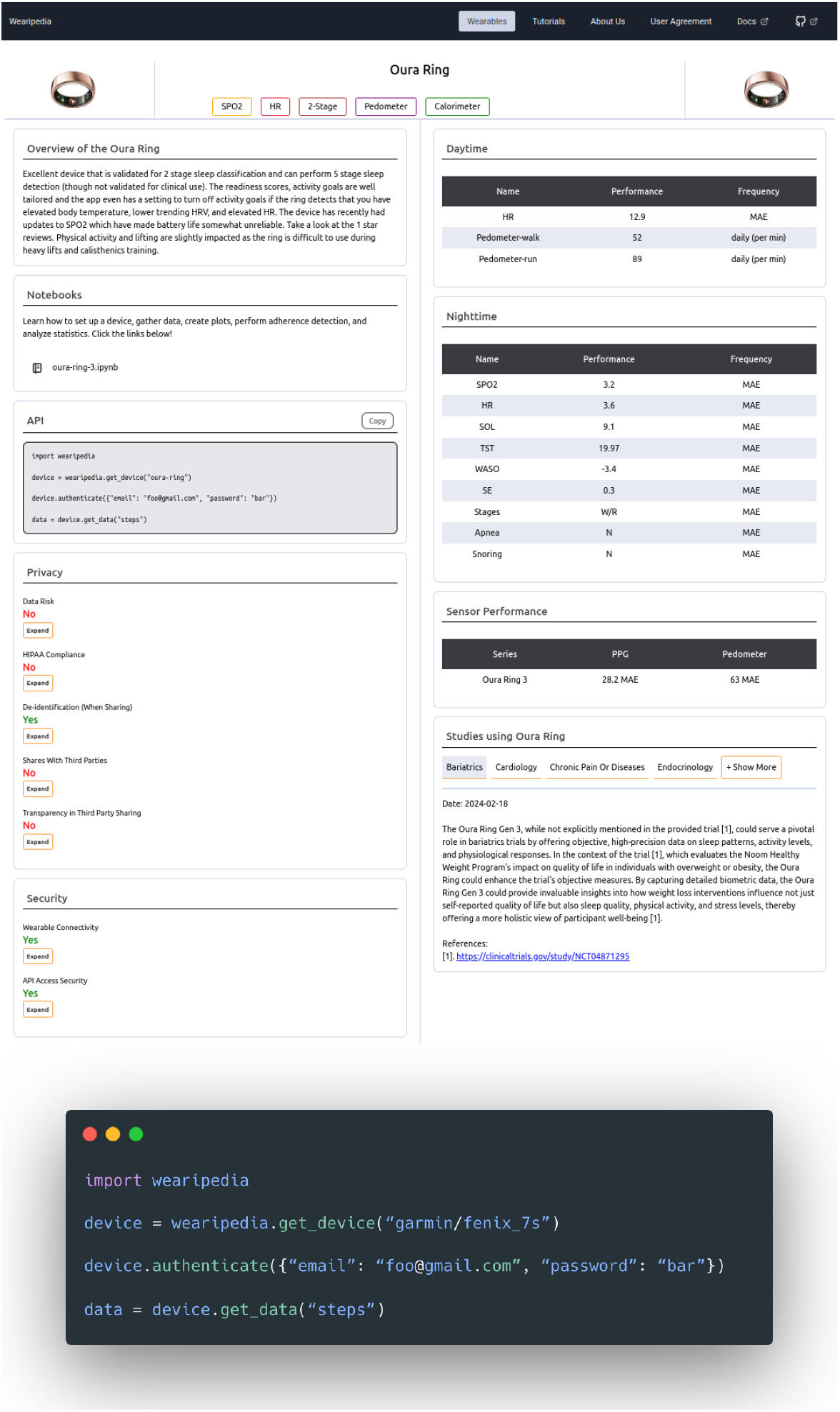
**Top:** Webpage of wearable device - Oura ring. Top left is an overview summary of the clinical performance and usability. Bottom left is security and privacy concerns with detailed drop-out descriptions. Right is clinical performance with links to studies. Performance metrics are in standardized formats detailed in a performance metric section, and reviewed in the top-left overview summary. Bottom right are clinicaltrials.gov studies that utilizes the wearable with autogenerated summaries by ChatGPT. **Bottom:** Code snippet demonstrating data extraction for the Garmin Fenix 7S device. It’s only four lines.

**Figure 2:**
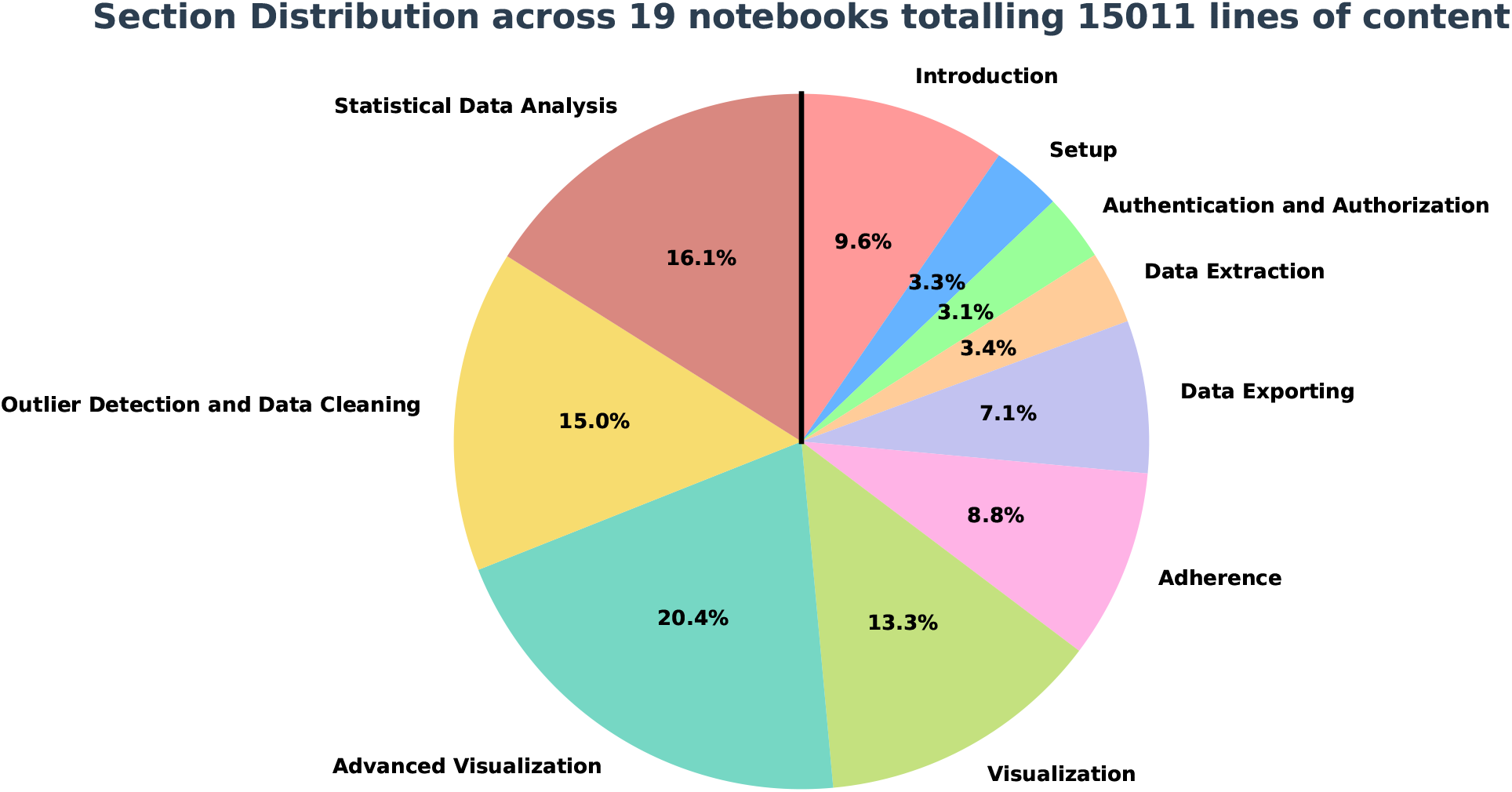
Notebook content distribution. the Wearipedia contains 21 notebooks: 19 single-participant data flow and 2 multi-participant data flows. This figure highlights the distribution of educational material over the 10 single-participant notebooks totaling 15,000 lines of code and text descriptions. Noticeable is that more than half of the content is catered towards more advanced data modeling (Advanced Visualization, Outlier Detection & Data Cleaning, and Statistical Data Analysis). This highlights a highly developed resource for clinical researchers to get started on their wearable data analysis journey.

The wearable tech market, valued at US$121.7 billion in 2021, is expected to surpass US$392.4 billion by 2030, driven by major tech companies entering healthcare. Five companies (Apple, Fitbit, Samsung, Xiaomi, Huawei) dominate 60% of the market, restricting data access and complicating clinical research due to proprietary systems and limited APIs. Research-oriented wearables offer better support but are expensive and less reliable. Moreover, there’s constant change with vendors abruptly discontinuing products and platforms.

We developed the Wearipedia, which is a comprehensive database covering 19 wearable devices and 5 apps in order to provide easy access for clinicians and researchers to downloading wearable data. Critically, it addresses privacy, usability, and data extraction challenges. The platform includes extensive Python resources, aiming to assist clinical researchers in selecting and using wearables effectively. Unlike existing databases, Wearipedia offers in-depth analysis and novel simulation tools—filling a gap in pre-clinical and operational wearable research. The database is open-source and continuously updated, providing valuable resources for both novice and experienced researchers and requires only basic Python programming skills. Specifically, our Python package, also called `wearipedia`, streamlines the process of extracting data from wearable device APIs. With just four lines of code, anyone is able to extract data from any wearable device included in our package, as seen in Figure 1.

#### Research in context

##### Evidence before this study

The Wearipedia ties together many novel topics into one database, including: privacy, security, data extraction, open source, educational material, clinical trial tracking. As the purpose of the Wearipedia is to tie together these resources to compare with public websites and databases.

Website and database: We performed a Pubmed search for ((“wearable” OR “wearables”) AND (“clinical research” OR “clinical studies” OR “healthcare” OR “biomedical research”) AND (“data studies” OR “healthcare” OR “biomedical research”) AND (“data extraction” OR “data access” OR “data management”) AND (“databases” OR “database”)) with 25 hits and a Google search for: “Wearable databases” in which we investigated the first 20 hits, and asked OpenAI’s GPT-4o for databases of wearables.

Our initial search revealed a patchwork of resources on wearable devices; databases like PhoneDB, GoHumanFirst, AtalasEDU, Vandrico, and DataverseNo^12^ providing only basic details such as price, production year, and sensor types.

##### Added value of this study

Website and database: To our knowledge, this is the first study to provide an open-source unified comprehension of wearable devices. We scrutinized privacy & security issues while offering detailed, device-specific coding resources for data extraction and analysis. By integrating aforementioned resources, we provide a holistic overview of each device in just one page. This sets a new standard for how researchers interact with wearable technology.

##### Implications of all the available evidence

Navigating the complexities of wearable tech is no small feat. With Wearipedia, clinical researchers of all levels have a single, comprehensive hub that makes wearable device selection and use both straightforward and effective, accelerating advances in decentralized clinical studies.

## Methods

To be adopted in the Wearipedia database and displayed on the website a host of content have to be covered in detail guiding users from search phase through deployment; Base requirements, API access & Wearipedia python package, Educational Material, Clinical Trial Tracking, and Privacy & Security.

### Basic device or app requirements

The mission is to empower clinical researchers performing decentralized clinical studies and trials with minimal participant burden. Consequently, the device/app must be wearable or usable at-home. It must be non-invasive, which for now excludes at-home blood testing. No in-clinic visits should be necessary. It must be related to some type of physiology measurement. A device must be purchasable at minimum in the US or EU, either publicly or through medical offices for patient treatment (i.e. certain continuous glucose monitoring (CGM) or heart monitoring devices).

### API access & Wearipedia python package

At the Wearipedia project, we commit to empowering clinical researchers by fully open-sourcing all device connectors. Therefore, any device included MUST have API access to device data. Devices without API access are excluded. Partial data is accepted (e.g. most devices give device summaries only). The device must be submitted to the open-source Wearipedia Python Package. In particular, we require that reasonable functionality is present and data simulators are built for allowing researchers with limited device access the ability to test devices before purchasing them. Moreover, simulated data is necessary for developed educational content. Devices will be reviewed on a device-by-device basis. Details on device submission processes can be found in Appendix A.

### Educational Material

To strengthen the clinical research community, we developed significant educational resources for clinical researchers with limited exposure to programming and data analysis. In particular, we want to guide clinical researchers through the basics of conducting and evaluating data from wearable devices. Our format of choice is Jupyter Notebooks in the Python coding language. For each educational notebook, we require the following steps for a notebook covering a single wearable with a single participant and a single study coordinator.

1. A 1-page letter detailing setup for the user, and a formal guide of what is expected of the user in the remainder of the clinical study.
2. A setup guide for the clinical researcher to ensure access to data for one user.
3. Data extraction using the Wearipedia python package for one user.
4. Data simulation using the Wearipedia python package
5. Guide on how to port data to R, Matlab, Excel, and standard data formats using the Wearipedia python package.
6. Minimum 2 plots visualizing details of wearable data
7. Guide, and visualization, on how to remove non-adherence for a custom period and custom adherence percentage (e.g. days, weeks, months).
8. Guide, and visualization, on how to remove outliers using statistical testing.
9. Guide, and visualization, on how to test a hypothesis.

The notebook must be subsequently submitted to the repository, and accepted through internal code-review ensuring it meets strict educational standards.

### Clinical Trial Tracking

Any device must have automatically updated summaries from the newest data available about the device on clinicaltrials.gov. In particular, we utilize the CliniDigest^13^ tool that scrapes, filters, and classifies clinical trials for wearable devices across 14 medical categories. Setting up CliniDigest for a new device is easy and simply requires defining useful keywords that sufficiently capture the clinical trials of choice. Details on what keywords to use, examples of other devices, and formal requirements for making a pull request is available at https://github.com/Stanford-Health/CliniDigest. The CliniDigest tool is automatically run every Sunday and updates summaries if new trials are available.

### Privacy & Security Device Evaluation

Privacy and security safeguards grow ever more urgent as digital systems erode individual control over personal data. While scholars still debate an exact definition of privacy^14–19^, most agree privacy underpins autonomy and liberty and thus warrants strong regulation^20^. Additionally, consentless sharing of data, real-time tracking of sensitive information, ambiguous terms of services, and weak authentication or encryption protocols are widespread^21–24^. The World Economic Forum lists data fraud and cyber attacks among the five likeliest global threats this decade^25^, and wearable ecosystems supply fertile ground—limited processing power, small batteries, unsecured Bluetooth, and rushed design cycles^26–29^. Lax developer practices^30^ and documented hacks^31^ have already exposed user data. Roughly one-third of U.S. adults already use a wearable device^32–34^ with sensors that record highly confidential health metrics^35,36^, yet are unregulated by the Food and Drug Administration and is beyond comprehensive laws such as HIPAA^37–39^. API-centric architectures enlarge the attack surface—public endpoints and microservices enable scraping or credential-stuffing, while insecure companion apps spread vulnerabilities from wrist to network^40–43^.

Scholars therefore urge a dedicated, binding privacy standard for health-monitoring wearables. In particular, we designed a set of relevant metrics to rate the privacy and security of wearable devices, documented in full in Appendix D. The privacy and security rating of each wearable device is measured under seven different criteria (5 under Privacy, 2 under Security) in Table 1.

**Table 1:** Brief descriptions of all seven privacy and security rating metrics. 5 ratings are under privacy, with 2 others under security. Longer, in-depth descriptions of each rating is provided in Appendix D.

| Metric Category | Rating | Brief Description |
| --- | --- | --- |
| Privacy | Low data risk | Wearable collects potentially sensitive information |
| Privacy | HIPAA compliant | Company does not collect personally identifiable information defined under HIPAA |
| Privacy | De-identification when sharing | Company de-identifies data when publicizing or sharing with other entities |
| Privacy | No third party sharing | Company commits to not sharing with third-parties |
| Privacy | Transparency in third party sharing | Company is exhaustively transparent with which entities data may be shared to |
| Security | Secure wearable connectivity | Wearable utilizes WiFi to connect to persistent stores |
| Security | Secure API access | Wearable provides API access through OAuth 2.0 protocol |

### Wearipedia Website and Database

We recognize that the information presented in the Wearipedia database can be overwhelming. Wearable biomedical sensors have significantly changed the landscape of clinical research and the requirements for clinical researchers. In particular, with understanding minute details on complex electrical devices and convoluted coding projects. To ease the process from start to finish we have adopted modern user-experience practices in designing a website for searching through Wearipedia entries and visualizing an entry on a single simple and intuitive page. The website was developed in Javascript with MongoDB, Express.js, and Node.js, then hosted on AWS. Each Wearipedia database entry is stored as a JSON entry. The raw Wearipedia JSON entries are open-source. The website design and layout is kept private due to security concerns.

## Results

Following the requirements outlined in the methodology section, we have the following as per April 2025: 19 Wearables and 5 Apps; 7 smart watches; 4 fitness trackers; 1 chest straps; 2 CGM devices; 1 smart ring; 1 arm strap; 1 under the bed sleep tracker; 1 smart scale; 2 apps for diet tracking; 1 app for questionnaires; and 1 apps for data storage.

### API access & Wearipedia python package

In Table 2, we cover API type, sampling rates, data simulators, and API-related concerns along with required permissions. The sampling and concerns are of particular importance as while a device might potentially have the capability to empower certain studies, poor sampling or data access can make it infeasible in practice. Notably, Whoop strap only has daily summaries, Garmin Fenix uses Oauth 1.0 which is insecure, Oura ring and Fitbit requires team approval for full data access, and Apple Watch does not have a public API, but requires developing and proceeding an app for data access. Interestingly, apps such as EliteHRV have developed a niche product to interface with Polar for high resolution, beat-to-beat data.

**Table 2:**
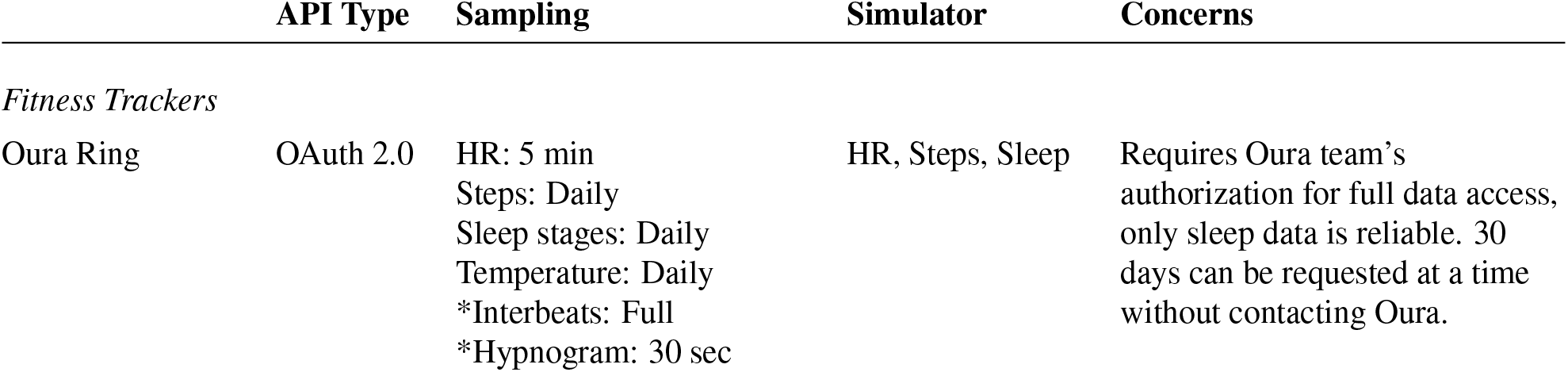
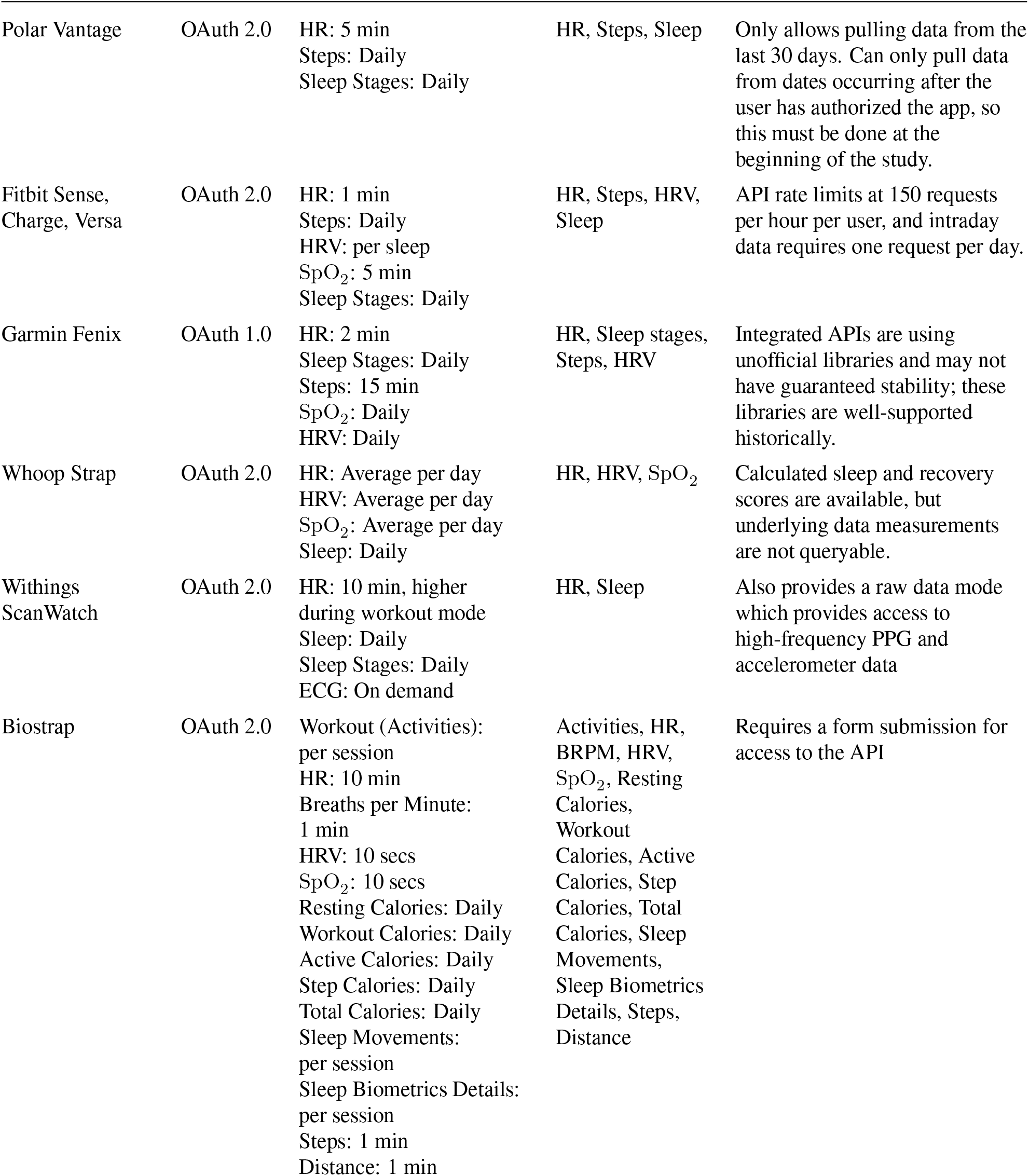
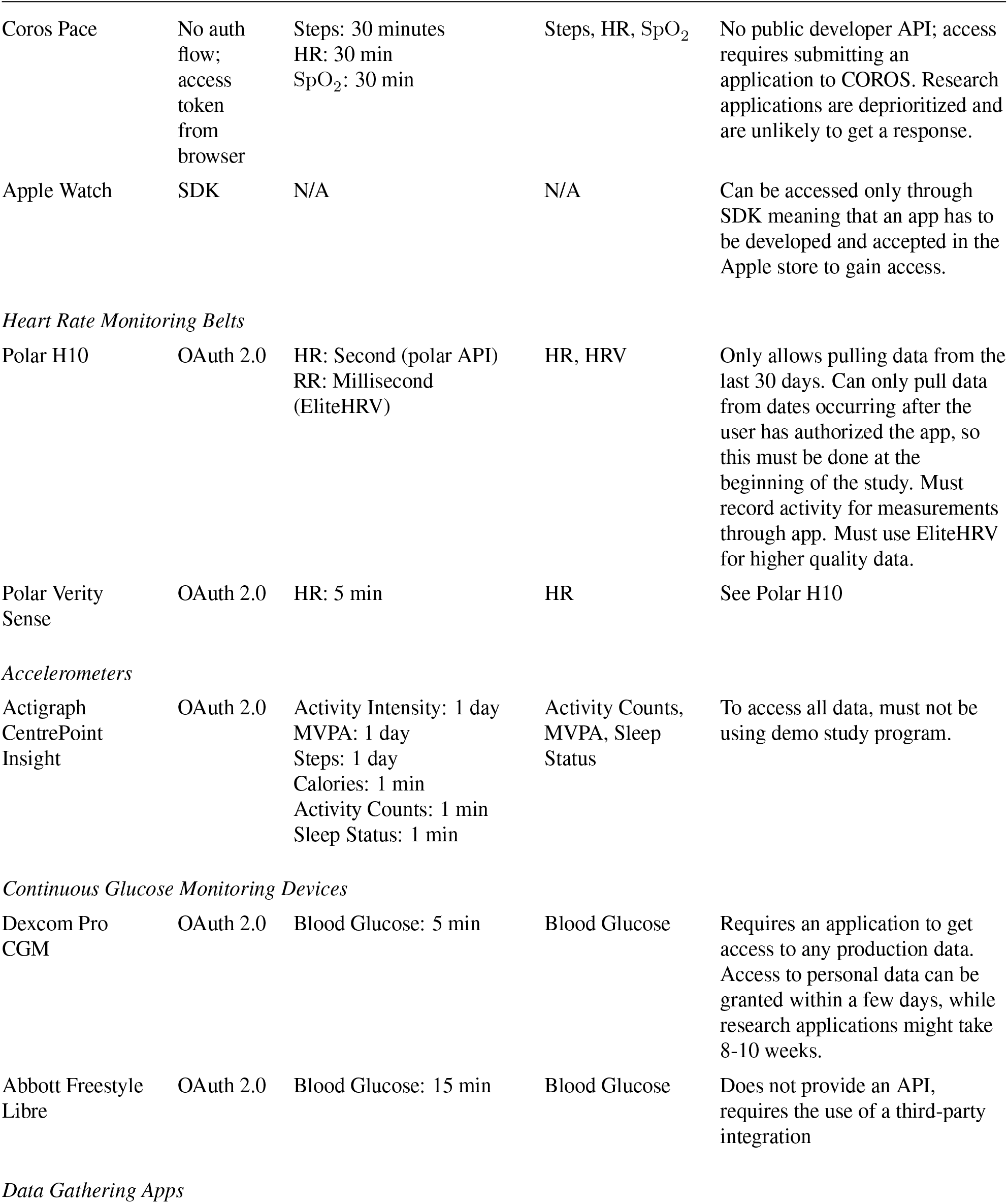
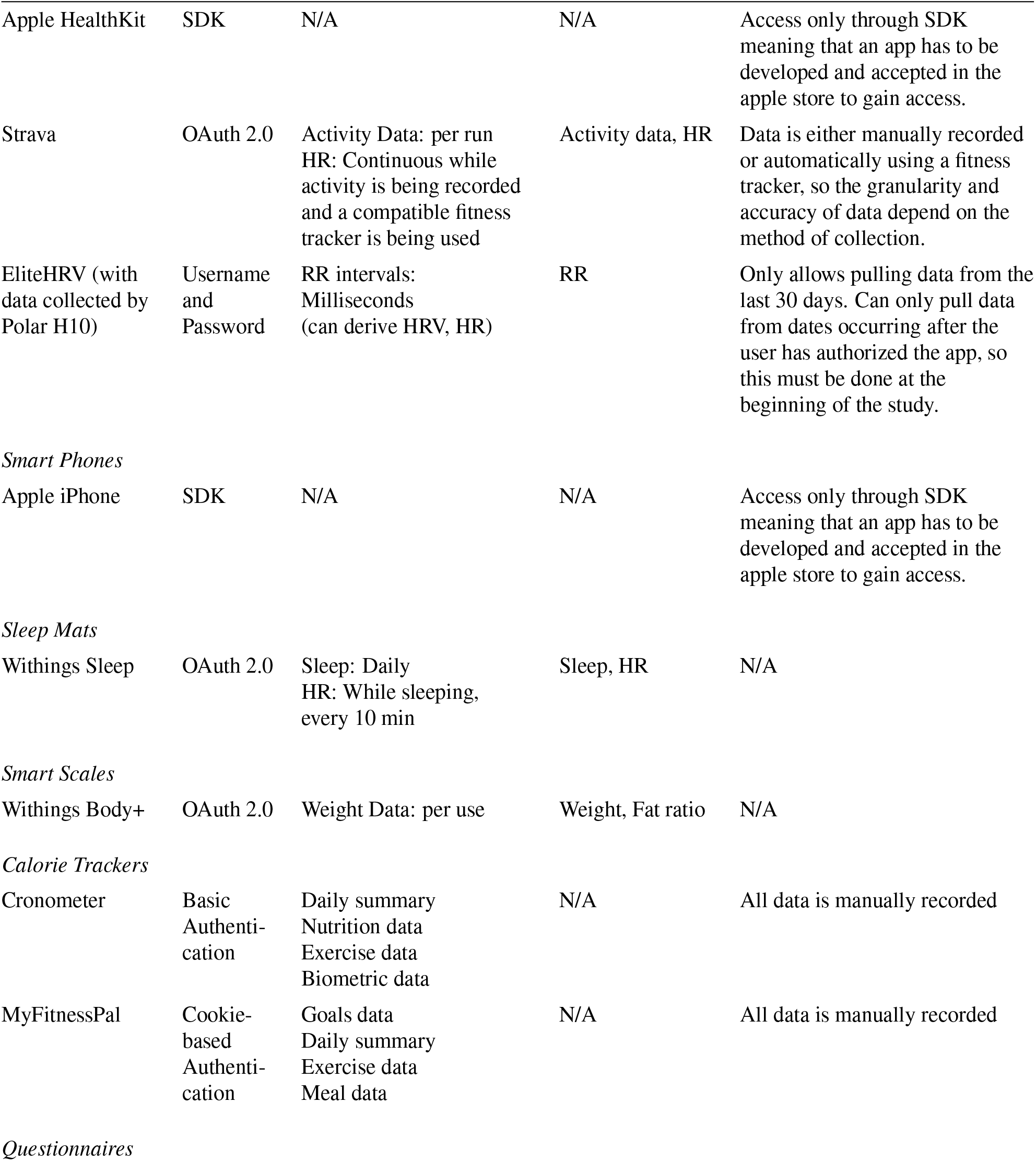
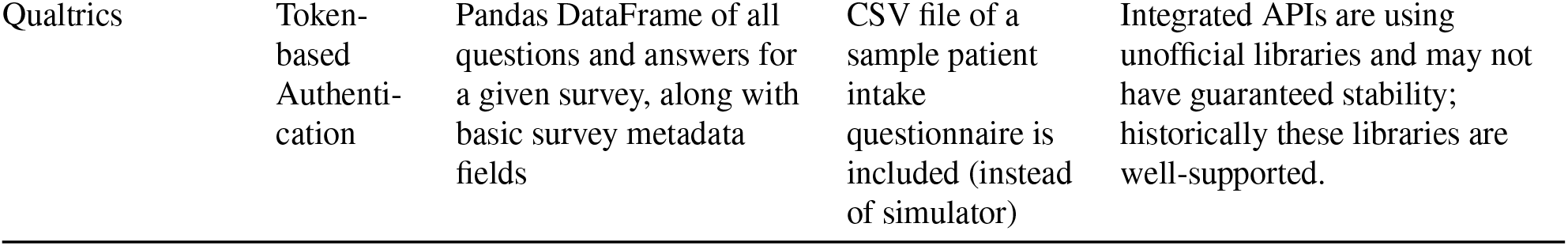
Information on API type, sampling details, simulator, and potential concerns for each wearable device sectioned by fitness trackers, HR belts, accelerometers, CGM devices, data gathering apps, smart phones, sleep mats, smart scales, calorie trackers, and questionnaires. *: includes special deals with the wearable vendor such as Fitbit Intraday and Oura Labs

|  | API Type | Sampling | Simulator | Concerns |
| --- | --- | --- | --- | --- |
| *Fitness Trackers* |  |  |  |  |
| Oura Ring | OAuth 2.0 | HR: 5 min<br>Steps: Daily<br>Sleep stages: Daily<br>Temperature: Daily<br>*Interbeats: Full<br>*Hypnogram: 30 sec | HR, Steps, Sleep | Requires Oura team's authorization for full data access, only sleep data is reliable. 30 days can be requested at a time without contacting Oura. |
| Polar Vantage | OAuth 2.0 | HR: 5 min<br>Steps: Daily<br>Sleep Stages: Daily | HR, Steps, Sleep | Only allows pulling data from the last 30 days. Can only pull data from dates occurring after the user has authorized the app, so this must be done at the beginning of the study. |
| Fitbit Sense, Charge, Versa | OAuth 2.0 | HR: 1 min<br>Steps: Daily<br>HRV: per sleep<br>SpO<sub>2</sub>: 5 min<br>Sleep Stages: Daily | HR, Steps, HRV, Sleep | API rate limits at 150 requests per hour per user, and intraday data requires one request per day. |
| Garmin Fenix | OAuth 1.0 | HR: 2 min<br>Sleep Stages: Daily<br>Steps: 15 min<br>SpO<sub>2</sub>: Daily<br>HRV: Daily | HR, Sleep stages, Steps, HRV | Integrated APIs are using unofficial libraries and may not have guaranteed stability; these libraries are well-supported historically. |
| Whoop Strap | OAuth 2.0 | HR: Average per day<br>HRV: Average per day<br>SpO<sub>2</sub>: Average per day<br>Sleep: Daily | HR, HRV, SpO<sub>2</sub> | Calculated sleep and recovery scores are available, but underlying data measurements are not queryable. |
| Withings ScanWatch | OAuth 2.0 | HR: 10 min, higher during workout mode<br>Sleep: Daily<br>Sleep Stages: Daily<br>ECG: On demand | HR, Sleep | Also provides a raw data mode which provides access to high-frequency PPG and accelerometer data |
| Biostrap | OAuth 2.0 | Workout (Activities): per session<br>HR: 10 min<br>Breaths per Minute: 1 min<br>HRV: 10 secs<br>SpO<sub>2</sub>: 10 secs<br>Resting Calories: Daily<br>Workout Calories: Daily<br>Active Calories: Daily<br>Step Calories: Daily<br>Total Calories: Daily<br>Sleep Movements: per session<br>Sleep Biometrics Details: per session<br>Steps: 1 min<br>Distance: 1 min | Activities, HR, BRPM, HRV, SpO<sub>2</sub>, Resting Calories, Workout Calories, Active Calories, Step Calories, Total Calories, Sleep Movements, Sleep Biometrics Details, Steps, Distance | Requires a form submission for access to the API |
| Coros Pace | No auth flow; access token from browser | Steps: 30 minutes<br>HR: 30 min<br>SpO<sub>2</sub>: 30 min | Steps, HR, SpO<sub>2</sub> | No public developer API; access requires submitting an application to COROS. Research applications are deprioritized and are unlikely to get a response. |
| Apple Watch | SDK | N/A | N/A | Can be accessed only through SDK meaning that an app has to be developed and accepted in the Apple store to gain access. |
| *Heart Rate Monitoring Belts* |  |  |  |  |
| Polar H10 | OAuth 2.0 | HR: Second (polar API)<br>RR: Millisecond (EliteHRV) | HR, HRV | Only allows pulling data from the last 30 days. Can only pull data from dates occurring after the user has authorized the app, so this must be done at the beginning of the study. Must record activity for measurements through app. Must use EliteHRV for higher quality data. |
| Polar Verity Sense | OAuth 2.0 | HR: 5 min | HR | See Polar H10 |
| *Accelerometers* |  |  |  |  |
| Actigraph<br>CentrePoint<br>Insight | OAuth 2.0 | Activity Intensity: 1 day<br>MVPA: 1 day<br>Steps: 1 day<br>Calories: 1 min<br>Activity Counts: 1 min<br>Sleep Status: 1 min | Activity Counts, MVPA, Sleep Status | To access all data, must not be using demo study program. |
| *Continuous Glucose Monitoring Devices* |  |  |  |  |
| Dexcom Pro CGM | OAuth 2.0 | Blood Glucose: 5 min | Blood Glucose | Requires an application to get access to any production data. Access to personal data can be granted within a few days, while research applications might take 8-10 weeks. |
| Abbott Freestyle Libre | OAuth 2.0 | Blood Glucose: 15 min | Blood Glucose | Does not provide an API, requires the use of a third-party integration |

|  | **API Type** | **Sampling** | **Simulator** | **Concerns** |
| --- | --- | --- | --- | --- |
| Apple HealthKit | SDK | N/A | N/A | Access only through SDK meaning that an app has to be developed and accepted in the apple store to gain access. |
| Strava | OAuth 2.0 | Activity Data: per run<br>HR: Continuous while activity is being recorded and a compatible fitness tracker is being used | Activity data, HR | Data is either manually recorded or automatically using a fitness tracker, so the granularity and accuracy of data depend on the method of collection. |
| EliteHRV (with data collected by Polar H10) | Username and Password | RR intervals: Milliseconds (can derive HRV, HR) | RR | Only allows pulling data from the last 30 days. Can only pull data from dates occurring after the user has authorized the app, so this must be done at the beginning of the study. |
| *Smart Phones* |  |  |  |  |
| Apple iPhone | SDK | N/A | N/A | Access only through SDK meaning that an app has to be developed and accepted in the apple store to gain access. |
| *Sleep Mats* |  |  |  |  |
| Withings Sleep | OAuth 2.0 | Sleep: Daily<br>HR: While sleeping, every 10 min | Sleep, HR | N/A |
| *Smart Scales* |  |  |  |  |
| Withings Body+ | OAuth 2.0 | Weight Data: per use | Weight, Fat ratio | N/A |
| *Calorie Trackers* |  |  |  |  |
| Cronometer | Basic Authentication | Daily summary<br>Nutrition data<br>Exercise data<br>Biometric data | N/A | All data is manually recorded |
| MyFitnessPal | Cookie-based Authentication | Goals data<br>Daily summary<br>Exercise data<br>Meal data | N/A | All data is manually recorded |

Table 2: Information on API type, sampling details, simulator, and potential concerns for each wearable device sectioned by fitness trackers, HR belts, accelerometers, CGM devices, data gathering apps, smart phones, sleep mats, smart scales, calorie trackers, and questionnaires.
|  |  |  |  |  |
| --- | --- | --- | --- | --- |
| Qualtrics | Token-based Authentication | Pandas DataFrame of all questions and answers for a given survey, along with basic survey metadata fields | CSV file of a sample patient intake questionnaire is included (instead of simulator) | Integrated APIs are using unofficial libraries and may not have guaranteed stability; historically these libraries are well-supported. |

### Educational Material

A major contribution of the Wearipedia is helping clinical researchers adopt python and guide data extraction, visualization, and analysis pipelines. Through detailed python notebooks, broken into 9 topics as covered in Figure **??**, we provide more than 15,000 lines of code and text description.

### Clinical Trial Tracking

A clear understanding of previously conducted research is paramount to choosing the best device and planning a new clinical trial. CliniDigest provides summaries of clinical trials involving wearable devices within 14 different fields of medical specializations. Updated automatically weekly through data extraction from ClinicalTrials.gov, CliniDigest produces and publishes new summaries encapsulating the current state of research using wearable devices. The pipeline includes papers up to five years old and a list of regular expressions encapsulating the search terms defined in Appendix C. A wide range of wearable devices are used in clinical trials, as depicted in Figure 3. Devices such as Abbott Freestyle Libre and Dexcom G Pro are primarily used for one medical field. In contrast, other devices, such as the iPhone and Fitbit devices, are used across several medical fields. As designed, as the number of clinical trials being summarized in a given summary increases, the summary becomes more general, citing fewer trials while still encompassing the trends and uses of the given wearable device. For example, 120 clinical trials address endocrinology using the Abbot Freestyle Libre system, with only four specifically cited to provide concrete examples in the summary below.

**Figure 3:**
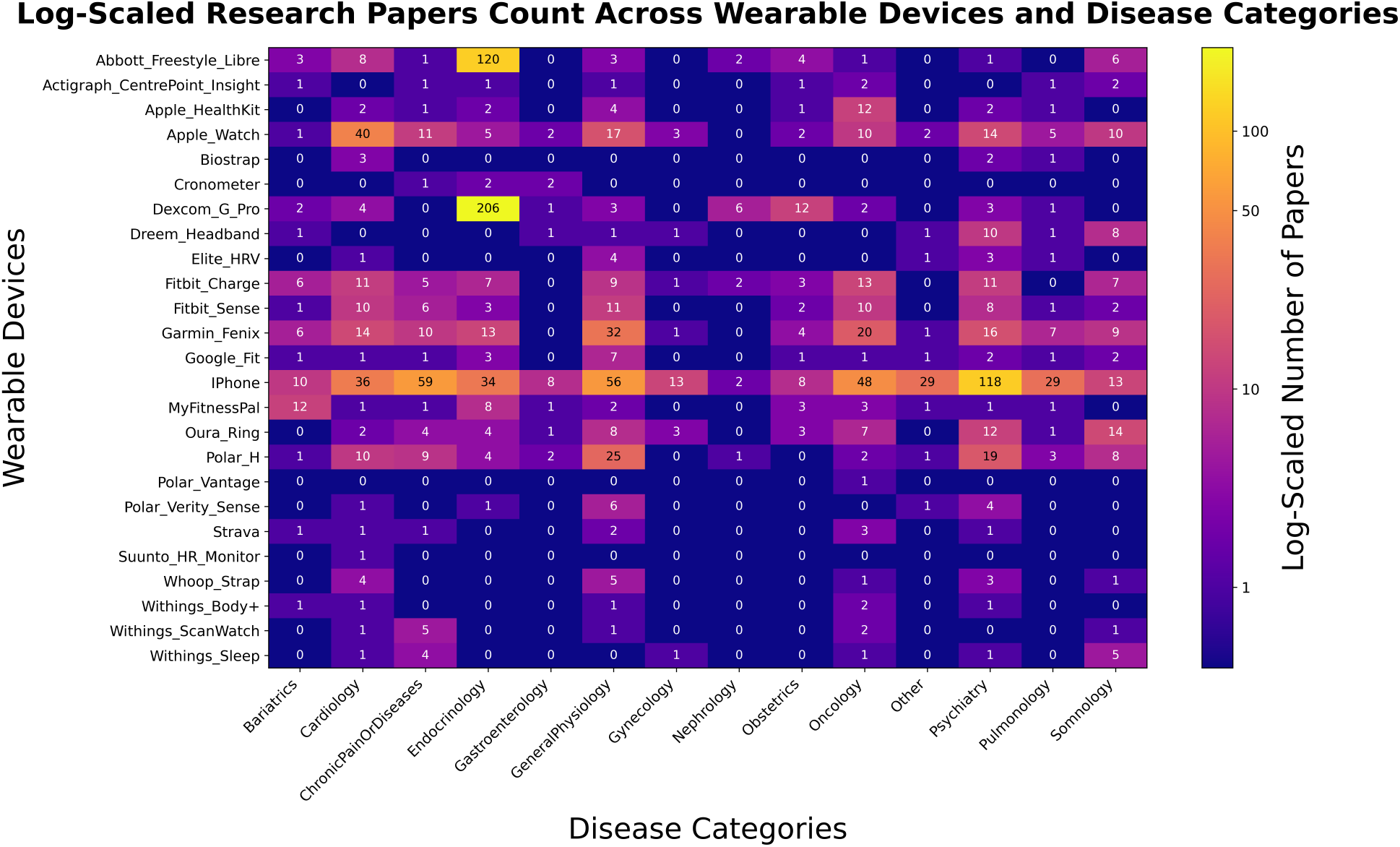
Clinical Trial count. y-axis is wearable device and x-axis is medical field. Each cube corresponds to the number of clinical trials on clinicaltrials.org that CliniDigest has found through its search criteria. These Clinical Trials are then included in a 200 word summary placed on each wearable device page on the Wearipedia website.

### Privacy & Security Device Evaluation

Each wearable was evaluated separately, but many had overlapping sources and gray literature that often resulted in identical scores. A full list of wearable devices are available in Table 3, denoted by company and model name. All wearables failed low data risk. By nature of the data collected, all wearables collected some data that could be categorized as a risk to privacy—either through primary or predictive data. All failed to be HIPAA compliant, as all vendors enforced some mechanism to collect personally identifiable information (PII) from users. This is most common in the form of name and e-mail address, though quite a few other vendors collected other PII. No company offered full access to their wearable device without requiring users to relinquish PII. Some wearable devices included a clause within their privacy policy or terms of service about de-identifying the data when sharing with other parties. Those that did not include this clause were Abbott Libre 2, Actigraph CPI, Coros Pace 2, Dexcom Pro CGM, Fitbit Sense/Charge 4, Garmin Fenix 7S, Nutrisense CGM Patch, Oura Ring 3, Polar H10/Vantage 2/Verity Sense, SleepOn go2sleep, Suunto HR Monitor, and Withings Body+/ScanWatch/Sleep. It may be the case that these wearables deidentify their data when sharing with other parties, but there was no explicit mention within their terms of use or privacy policies. All wearables included a clause about sharing data with third parties. This was most commonly stated in the privacy policy or terms of use. Similarly, all but one did not exhaustively disclose the list of third parties that they may share data with. Most wearables stated some variation of “other”, “miscellaneous”, or “various” third parties that may currently or at a different time point gain access to user data. Only Garmin exhaustively listed the third parties that will gain access to user data.

**Table 3:** Wearable devices and smartphone apps evaluated for privacy and security concerns across seven different categories. A green check mark signifies a satisfactory result defined by a specified threshold, whereas a red x mark means it is unsatisfactory. A dashed line indicates that result is not applicable (e.g. Apple health kit is not a wearable), so secure wearable connectivity cannot be described. Detailed descriptions of each metric as well as the threshold for satisfactory performance are detailed in Appendix D. Citations for how each rating was achieved is also provided in D.4.

| Company | Model Name | Low Data Risk | Compliant with HIPAA | De-identifies Data When Sharing | No Third Party Sharing | Transparency in Third Party Sharing | Secure Wearable Connectivity | Secure API Access |
| --- | --- | --- | --- | --- | --- | --- | --- | --- |
| Abbott | Libre 2 | ✗ | ✗ | ✗ | ✗ | ✗ | ✓ | — |
| Actigraph | CPI | ✗ | ✗ | ✗ | ✗ | ✗ | ✗ | ✓ |
| Apple | Health kit | ✗ | ✗ | ✓ | ✗ | ✗ | — | ✓ |
| Apple | Watch 5 | ✗ | ✗ | ✓ | ✗ | ✗ | ✓ | ✓ |
| Apple | Watch Ultra | ✗ | ✗ | ✓ | ✗ | ✗ | ✓ | ✓ |
| Apple | iPhone | ✗ | ✗ | ✓ | ✗ | ✗ | ✓ | — |
| Coros | Pace 2 | ✗ | ✗ | ✗ | ✗ | ✗ | ✗ | — |
| Dexcom | Pro CGM | ✗ | ✗ | ✗ | ✗ | ✗ | ✗ | ✓ |
| Fitbit | Sense | ✗ | ✗ | ✗ | ✗ | ✗ | ✓ | ✓ |
| Fitbit | Charge 4 | ✗ | ✗ | ✗ | ✗ | ✗ | ✓ | ✓ |
| Garmin | Fenix 7S | ✗ | ✗ | ✗ | ✗ | ✓ | ✓ | ✗ |
| Nutrisense | CGM patch | ✗ | ✗ | ✗ | ✗ | ✗ | ✓ | — |
| Oura | Ring 3 | ✗ | ✗ | ✗ | ✗ | ✗ | ✓ | ✓ |
| Polar | H10 | ✗ | ✗ | ✗ | ✗ | ✗ | ✓ | ✓ |
| Polar | Vantage 2 | ✗ | ✗ | ✗ | ✗ | ✗ | ✗ | ✓ |
| Polar | Verity Sense | ✗ | ✗ | ✗ | ✗ | ✗ | ✓ | ✓ |
| SleepOn | go2sleep | ✗ | ✗ | ✗ | ✗ | ✗ | ✗ | — |
| Suunto | HR Monitor | ✗ | ✗ | ✗ | ✗ | ✗ | ✗ | ✓ |
| Whoop | Strap 4.0 | ✗ | ✗ | ✓ | ✗ | ✗ | ✗ | ✓ |
| Withings | Body+ | ✗ | ✗ | ✗ | ✗ | ✗ | ✓ | ✓ |
| Withings | ScanWatch | ✗ | ✗ | ✗ | ✗ | ✗ | ✗ | ✓ |
| Withings | Sleep | ✗ | ✗ | ✗ | ✗ | ✗ | ✓ | ✓ |
| - | Cronometer | ✗ | ✗ | ✓ | ✗ | ✗ | — | — |
| - | MyFitnessPal | ✗ | ✗ | ✓ | ✗ | ✗ | — | ✓ |
| - | Strava | ✗ | ✗ | ✓ | ✗ | ✗ | — | ✓ |

## Discussion

### Most noticeable devices

#### Fitness

The Polar H10 is a chest strap that is considered the gold standard for many studies w.r.t. accurate heart rate and HRV measurements at only US$105 (excluding taxes) and long battery life. However, an additional app, EliteHRV is needed to extract data at US$30-100 per month for a cohort. Moreover, a belt could lower user adherence compared to a ring or wrist strap.

#### Sleep

We find that the Oura ring is a great option for sleep study tracking due to great performance on sleep metrics, high quality data access, long battery life, and low concerns with user compliance due to the unobstructedness of wearing the device. However, the Oura ring is pricey (US$349 excluding taxes), needs special access for high quality data extraction, demonstrates low capabilities in step and physical activity detection, and potentially requires removal before manual labor or strength training—resulting in the loss of valuable data during such activities.

#### Budget friendly

The Fitbit Charge 6 costs only US$160 (excluding taxes), has medium-to-high sleep and step prediction capability, battery life, high quality data access, can be used both while sleeping and for most physical activity.

#### Apple Watch

While achieving high performance across the board, the Apple Watch has concerns with battery life, being uncomfortable while sleeping, a large screen that can interfere with study participants, and limited access to high quality data. For improved battery life Apple Ultra can be considered, but it comes at a hefty price point.

### Interoperability between devices

Polar for fitness activities, Fitbit for step counts, Oura for sleep, Dexcom for CGM, and MyFitnessPal for diet tracking—should be simple? Unfortunately, no vendor has kept interoperability in mind and merging datasets as well as timestamp is the clinical study coordinators’ responsibility. The Wearipedia python package and the educational materials offer tools to ease the process (how to extract, find missing data, visualize, and perform statistical analysis). However, there is limited merging material.

### Wearipedia website and Database

The Wearipedia project provides curated information about wearable devices to accelerate pre-clinical studies and education of wearable devices. The website is easy to navigate and well documented. Currently 24 wearable devices and 5 apps are covered. The educational material is meant as an inspiration, and a starting point, for clinical researchers looking to work with wearable devices. Given all elements of the Wearipedia project is open source, future additions of wearable device and app pages from both in-house and external contributors is supported and encouraged. In the future we would like to extend to actively support clinical studies from start to finish.

### What wearables devices to include next?

#### Popular fitness trackers

We have left out some key commercially, and cheap, wearables devices. Some do not have easily accessible data streams, including: Samsung Galaxy Watch, Xiaomi Mi Band, AmazFit, Suunto, and Coros. Some can have data partially accessed through the Apple HealthKit or Strava, but in that case we already have them covered.

#### EEG headbands

We covered the Dreem Headband 3 in detail, but as the company was acquired and discontinued their helmet we have excluded it from our analysis. Other popular high-quality headbands include Hypnodyne Zmax and OpenBCI, these are quite interesting but also very expensive.

#### Raw data access

Almost no commercially available device provides raw data access, Fitbit SDK gives some limited accelerometer/gyroscope for apps. OpenBCI offers EEG and PPG sensors with full access to sensors, which we have note covered yet. Other research oriented wearable devices might do the same

#### Research wearable devices

We have tried to collaborate with wearable devices specifically designed for researchers, but it is cumbersome to access a single device for our purpose. Hopefully materializing this public platform allows research device companies to see the value and provide their device and API connectors to our open-source platform.

### Use in clinical research

To run a distributed clinical study with wearables HIPAA compliance and a data risk assessment is required. The Wearipedia python package simplifies communication with wearable vendor. However, it is not a end-to-end solution. Wearable vendors have to be approved through individual data risk assessment. The Wearipedia python package has to be stalled on a secure server and pipe the data into a secure and HIPAA approved database.

### Human assays

For now, we have chosen to not pursue any human assays, even though in particular semen analysis is very easy to procure and well tested by now.

## Data Availability

There is no data generated, but all code is open-source and publicly available

https://wearipedia.com

https://github.com/Stanford-Health/wearipedia

https://github.com/Stanford-Health/wearable-notebooks

## Ethics in research

We have seen that wearable devices offer unprecedented opportunities to capture real-time, high-quality data in decentralized clinical trials (DCTs) at little cost and limited patient burden. However, the use of wearables in DCTs also poses critical challenges that must be addressed to protect the rights and well-being of participants. a) Including minorities is pivotal in the early phases, in particularly as the algorithms and methodologies developed in these early stages need to be as inclusive as possible, i.e. is a US$800 Garmin watch necessary, or would a US$100 Fitbit do just as well? Moreover, parameters such as melanin impacts PPG readings, and digital-readiness is not a given^44,45^. b) Privacy concerns escalate when continuous data streams from wearable devices are transmitted via multiple platforms, in particular as most wearable vendors do not prohibit sharing data with unknown third-parties and dipping into on-device databases can be coerced without the participant realizing it. With easily derived features such as cycle, pregnancy status, heart issues, infectious disease status, and diabetes, it puts the participant at risk for partner violence, imprisonment under anti-abortion rulings, and being passed up for career opportunities or insurance company bias.^45,46^. c) As most health issues are faced by an aging population the customer user base is mainly composed of the younger population that has adopted wearable technology. This could lead to an exclusion of the population most in need for novel wearable solutions^47^. Conclusively, there’s a need for a development of standards to ensure minorities and elderly are well represented in DCTs, with the low cost and ease of deployment this should be easier than ever. The more challenging task is to ensure privacy for individuals, in particular women of reproductive age, but as the algorithms keep getting better, possibly everyone. Current solutions are unfortunately quite limited as this would remove most commercially available products.

## Data Sharing

Wearipedia python package: https://github.com/Stanford-Health/

Wearipedia website: https://wearipedia.com

CliniDigest: https://github.com/Stanford-Health/CliniDigest

Wearipedia data: https://github.com/Stanford-Health/wearipedia-data

## Declaration of interests

### Contributors

**Alexander Johansen**: Project lead, review of educational notebooks, website content

**Kyu Hur**: Website development, privacy and security section

**Jack Hung**: Wearipedia python package, notebooks: Polar H10/EliteHRV, Verity Sense, Abbott Freestyle Libre, Nutrisense, Actigraph CentrePoint Insight

**Rodrigo Castellon**: Wearipedia python package, Whoop Strap, Withings ScanWatch, Body+, Sleep, Dreem Head-band 3, Garmin Fenix, Dexcom CGM

**Tristan Peng**: Website development, CliniDigest

**Stephanie Ren**: Wearipedia python package, data extractors

**Renee White**: CliniDigest

**Chanyeong Park**: Platform for Clinical Researchers

**Allison Lau**: Fitbit, Garmin, Whoop Strap, Dexcom Pro CGM

**Saarth Shah**: Polar Vantage, Strava, Cronometer, MyFitnessPal

**Hee Jung Choi**: Biostrap, Qualtrics

**William Wang**: Whoop Strap, Withings ScanWatch, Body+, Sleep, Dreem Headband 3, Garmin Fenix, Dexcom CGM

**Pann Sripitak**: CliniDigest

**Mohamed Elhusinni**: Oura ring, Coros Pace

**Michael Snyder**: PI

### Conflicts of interests

MPS is a cofounder and scientific advisor of Crosshair Therapeutics, Exposomics, Filtricine, Fodsel, iollo, InVu Health, January AI, Marble Therapeutics, Mirvie, Next Thought AI, Orange Street Ventures, Personalis, Protos Biologics, Qbio, RTHM, SensOmics. MPS is a scientific advisor of Abbratech, Applied Cognition, Enovone, Jupiter Therapeutics, M3 Helium, Mitrix, Neuvivo, Onza, Sigil Biosciences, TranscribeGlass, WndrHLTH, Yuvan Research. MPS is a cofounder of NiMo Therapeutics. MPS is an investor and scientific advisor of R42 and Swaza. MPS is an investor in Repair Biotechnologies. All other authors declare no competing interests.

## Acknowledgments

This work was made possible by the support of the BV and Anu Jagadeesh Family Foundation. The authors would like to thank Helge Ræder for comments and review.

## Appendix A. Website and Database — Submission Walk-through

### Overview

To submit a device to the Wearipedia is a multistep process, first a device connector needs to be accepted, then an educational notebook, then CliniDigest and finally clinical and security evaluations. Device connector - The Wearipedia python package The wearipedia python package is hosted in an open-source Github project at https://github.com/stanford-health/wearipedia and details on submitting pull-requests to the repository are presented here. A functional data connector conveniently wraps the Application Programming Interface (API) or Software Development Kit (SDK) of the vendor to access data resulting in an easy-to-use interface.

A connector must have a simulator for key data types. This is to reduce unnecessary sharing of user credentials when building educational content and testing devices.

#### Educational notebook

The Wearipedia project has a set of educational open-source notebooks at https://github.com/Stanford-Health/wearable-notebooks. The notebooks are written in python and details how to extract and conduct clinical research using the wearable. To submit a notebook it should follow the general style of: https://github.com/Stanford-Health/wearable-notebooks/blob/main/notebooks/fitbit_charge_6.ipynb which includes: A 1-page letter detailing setup for the user, and a formal guide of what is expected of the user in the remainder of the clinical study. A setup guide for the clinical researcher to ensure access to data for one user. Data extraction using the Wearipedia python package for one user. Data simulation using the Wearipedia python package Guide on how to port data to R, Matlab, Excel, and standard data formats using the Wearipedia python package. Minimum 2 plots visualizing details of data Guide, and visualization, on how to remove non-adherence for a custom period and custom adherence percentage (e.g. days, weeks, months). Guide, and visualization, on how to remove outliers using statistical testing. Guide, and visualization, on how to test a hypothesis.

#### CliniDigest

CliniDigest provides overviews of wearable usage in current clinical studies sorted by medical fields. A natural language processing tool that extracts clinical trials from https://clinicaltrials.gov, then filters based on search terms (e.g. Fitbit), classifies across 14 clinical topics, and writes a summary using LLMs such as ChatGPT. To include a wearable the only requirement is to upload the additional search terms. Guidelines can be found at https://github.com/Stanford-Health/CliniDigest.

#### Privacy and Security

The Wearipedia provides a privacy and security evaluation of 25 wearable devices in the main text under Table 3. Guidelines can be found at https://github.com/Stanford-Health/wearipedia-data. The metrics are fully outlined under Appendix D and the source material for these evaluations can include the privacy policy, terms of service, or other gray literature provided by the company. To submit the privacy and security evaluations of a new device, a pull request can be opened according to the instructions outlined in the repository’s README.

## Appendix B. Wearipedia Python Package

Wearipedia is a Python package designed to standardize and simplify the process of interfacing with wearable devices. The package is built with modern software engineering practices and follows a modular, extensible architecture.

### Package Architecture

Wearipedia defines a device-agnostic abstraction layer that allows for easy integration of a new wearable device. The base of the package is the definition of the device interface, which defines the required methods that must be implemented by each new device. Each implementation follows the standardized interface while making device-specific optimizations.

One of the core pieces of each implementation is authentication. Because different devices use different protocols such as OAuth 2.0, OAuth 1.0, or basic authentication with username and password, each authentication per device type must be implemented separately with the right endpoints and parameters. For some devices, there is both an interactive and non-interactive version of authentication. The interactive version can be used in a user-friendly manner to prompt users through the authentication process, which may include clicking a link to the application and granting access. The non-interactive version can still be used to authenticate programmatically as needed.

On initialization, each device implementation must specify its valid data types for types of data that can be requested. It must also implement data access through a `get data` method. Each implementation should take in a standardized format of inputs such as `YYYY-MM-DD` dates as well as return outputs using standardized units. Devices are implemented to have both a real and synthetic data setting. The real setting queries for real user data and the synthetic setting generates fake data that can be used to develop data analysis code without accessing user data. Each device implementation should include tests for both the real and synthetic settings.

The package also includes a command line interface (CLI) mode which provides a user-friendly interface for interacting with data through Wearipedia.

### Appendix B.1. Technical Implementation Details

#### Appendix B.1.1. Development Environment

The package is supported for Python 3.9+ (except Garmin access, which is only supported in 3.10+) and follows modern development practices:

1. Dependency Management: Utilizes Poetry for dependency management and version control
2. Code Quality: Implements pre-commit hooks for code formatting and static analysis
3. Testing Framework: Comprehensive test suite using pytest, with coverage tracking
4. Documentation: Sphinx-based documentation system with ReadTheDocs integration

We chose to use Python for the development language due to its popularity for data analysis in research applications.

#### Appendix B.2. Design Decisions

##### Modularity

The package is designed with clear separation of concerns, allowing for:

1. Independent development of device drivers
2. Easy maintenance and updates

#### User Experience

1. Intuitive API design with consistent method naming
2. Clear parameter documentation
3. Type hints for better IDE integration
4. Comprehensive error messages
5. CLI interface
6. Wrappers around provided API functionality; e.g. combining multiple pages of data from paginated queries

## Appendix C. Clinical Trial Tracking

CliniDigest aims to provide clinical trial coordinators with easy and concise access to recent clinical trial developments. As such, it is imperative to use specific criteria for which clinical trials to include. Considering the date of the last update posted, enrollment count, and study status, the pipeline utilizes two complementary criteria for including a clinical trial. The criteria for a clinical trial to be included is as follows:

### Option 1: New trials

- Last Update Posted: 2 years ago — today
- Enrollment Count: ≥ 25
- Study Status (one of): Recruiting, Enrolling by Invitation, Active Not Recruiting, Not Yet Recruiting, Suspended, Completed

### Option 2: Current trials

- Last Update Posted: 5 years ago — 2 years ago
- Enrollment Count: ≥ 25
- Study Status (one of): Recruiting, Enrolling by Invitation, Active Not Recruiting

From the clinical trials that fit within the criteria to be searched, the regular expressions defined in the pipeline are checked for matches within each clinical trial’s data. For each medical device, it is necessary to identify the best search terms to identify clinical trials that use the given medical device. In order to do so, follow the steps detailed at https://github.com/Stanford-Health/CliniDigest. After identifying all clinical trials that use a wearable device, they are classified into fourteen medical fields. Each combination of a wearable device and medical combination with at least one clinical trial is then summarized using engineered prompts and OpenAI’s gpt-4-0125-preview model. The default summary length requested is 150-250 words. The summary length is shortened to 50-150 words if there are five or fewer clinical trials. Below are three examples: one with fewer than five trials, one with an average number of trials, and one with over two hundred trials.

> The purpose of a Polar H in pulmonology trials is to provide a reliable and non-invasive method for monitoring and assessing cardiovascular responses to various rehabilitation interventions in patients with respiratory conditions. By measuring heart rate variability and other hemodynamic responses, the Polar H facilitates the evaluation of the immediate effects of innovative exercise modalities, such as high-intensity interval exercises in children with asthma [1], and supports the safety and efficacy assessment of novel rehabilitation interventions like low-intensity exercise with blood flow restriction in COPD patients [2]. This technology is crucial for tailoring and optimizing pulmonary rehabilitation programs to enhance patient outcomes.

Example 1: Example of CliniDigest for the Polar H on Pulmonology. Out of 3 studies, 2 are selected and used for the summary.

> The Garmin Fenix, a sophisticated wearable technology, is increasingly being integrated into oncology trials to enhance patient monitoring, improve quality of life (QoL), and facilitate personalized interventions. Its purpose in these trials is multifaceted, primarily focusing on the collection of real-world data (RWD) to monitor physical activity, sleep quality, and heart rate, which are critical parameters in assessing the overall health and well-being of cancer patients [2][3]. For instance, in the REBECCA-2 study, the Garmin Fenix is utilized to gather RWD, enabling personalized follow-up care aimed at improving the QoL of breast cancer survivors by detecting signs of QoL deterioration and facilitating timely interventions [2]. Similarly, the eCAN JA project employs this device to automatically collect physical parameters, supporting tele-rehabilitation and tele-psychological support programs that aim to empower patients and improve their QoL through enhanced remote monitoring [3]. Moreover, the device’s role extends to pediatric oncology, where it aids in measuring recovery post-surgery through smartwatch data and questionnaires, highlighting its versatility across different age groups and cancer types [7]. The Garmin Fenix’s ability to seamlessly integrate into patients’ lives while providing valuable health metrics positions it as a pivotal tool in modern oncology trials, driving forward the paradigm of personalized and remote patient care.

Example 2: Example of CliniDigest for the Garmin Fenix on Oncology. Out of 20 studies, 3 are selected and used for the summary.

> The Dexcom G Pro, a continuous glucose monitoring (CGM) system, plays a pivotal role in advancing endocrinology trials by providing accurate, real-time glucose level data across various patient populations and conditions. Its purpose extends beyond mere glucose tracking; it serves as a critical tool in evaluating the efficacy and safety of diabetes management strategies, understanding the glycemic impact of pharmaceutical interventions, and exploring the physiological nuances of diabetes and its comorbidities. For instance, in trials evaluating the safety and effectiveness of automated insulin delivery systems in children and adults with type 1 diabetes, the Dexcom G Pro’s continuous glucose data is indispensable for assessing system performance in real-world settings [13][195]. Similarly, its use in studies exploring the impact of dietary interventions on glycemic control underscores its utility in non-pharmacological diabetes research [179]. Moreover, the Dexcom G Pro facilitates the investigation of glycemic variability and its clinical implications in conditions like polycystic ovary syndrome, thereby broadening our understanding of glucose dynamics beyond diabetes [170]. In the context of hospital care, its application in monitoring hospitalized patients with diabetes highlights its potential to improve inpatient glycemic management and reduce the risk of hypoglycemia [192]. Collectively, the Dexcom G Pro’s integration into endocrinology trials underscores its critical role in enhancing diabetes care through rigorous research, ultimately contributing to the development of personalized, data-driven treatment approaches that can significantly improve patient outcomes.

Example 3: Example of CliniDigest for the Dexcom G Pro on Endocrinology. Out of 206 studies, 5 are selected and used for the summary.

## Appendix D. Privacy and Security

Full descriptions of every metric under privacy and security provided in Table 1 are listed below. For each rating, a metric description elucidates the context and need for each rating, then a recommended performance which determines the threshold of whether the wearable passes or fails that category is described. 25 different wearables are graded on these 7 ratings in Table 3.

### Appendix D.1. Metric Description — Low Data Risk

Wearable devices collect highly sensitive personal data-sleep quality, activity levels, heart rate, glucose readings and location—which, if mismanaged, can cause serious harm to users^1–4^. Such primary data are straightforward to measure but demand strict controls. Beyond what is directly recorded, predictive data health risk scores, fitness estimates, lifestyle or behavioral patterns, mood or disease onset predictions can be inferred from primary signals. As inference methods advance, even innocuous measurements may reveal deeply personal insights^5–8^. This growing inferential power makes true anonymization impossible^9,10^. Some examples include body temperature for tracking period cycles or pregnancy^8^, heart rate for early COVID-19 detection^11^, and location for stalking or harassment^12–14^. Primary data categories commonly collected include physical activity (steps, distance, calories, pace, MET-minutes)^15,16^, physiological (heart rate variability, blood pressure, SpO_2_, skin temperature)^17^, sleep metrics (total sleep time, stages, interruptions, efficiency)^18^, and health status (glucose, hydration, BMI, menstrual tracking)^19,20^. Predictive data examples include health risk assessments (e.g. diabetes, cardiovascular disease)^21,22^, fitness metrics (VO_2_ max, recovery time)^23,24^, lifestyle and behavioral patterns (sleep habits, social interaction, sedentary time)^25,26^, and health insights (flu onset, mood, fatigue)^27^. Under privacy frameworks (HIPAA, GDPR), the highest risk categories are primary physiological, sleep and health status data, plus predictive health assessments and insights^28,29^. Misuse by insurers, employers or malicious actors can lead to discrimination, social stigma, psychological distress and financial loss^30,31^.

#### Recommended Performance

Wearable vendors should limit collection to only what is strictly necessary and implement robust safeguards across both primary and predictive data.

### Appendix D.2. Metric Description — Compliant with HIPAA

The Health Insurance Portability and Accountability Act (HIPAA), enacted in 1996, established comprehensive standards for protecting patient health information and was strengthened by the 2009 HITECH amendment to cover electronic health records, earning praise for its detailed privacy safeguards in practice^28,32,33^. As consumer wearables generate increasingly diverse and granular data streams, experts have called for HIPAA’s scope to expand to include sensor-derived information and for its provisions to be updated accordingly^34,35^. Under current rules, de-identification mandates suppression of 18 direct and indirect identifiers before any dataset release, and patient consent is required for disclosures of identifiable information^28^. Large scale anonymized repositories—such as NHANES—demonstrate how stripping these identifiers can enable valuable research while ostensibly preserving privacy^36^. However, numerous high-profile cases have shown that anonymized health data can be re-identified by linking with external sources: the Netflix Prize dataset was deanonymized through auxiliary movie ratings^37^, AOL search logs revealed user identities^38^, and medical records spurred the development of k-anonymity models after re-identification of supposedly private health data^39,40^. Wearable-derived datasets demonstrate volume, frequency, as well as richness and can face even greater re-identification risks^41,42^. To mitigate these vulnerabilities, wearable manufacturers should limit collection of HIPAA identifiers, apply robust de-identification techniques in line with HIPAA’s Privacy Rule, restrict data sharing to essential uses with explicit user consent, and acknowledge that no anonymization can be entirely foolproof.

#### Recommended Performance

Vendors should not collect any HIPAA identifiers.

Any data that falls under what is outlined as a HIPAA identifier would make a device non-HIPAA compliant. While most wearable devices are not covered under HIPAA, this law provides a comprehensive legal framework to determine what may be important identifying data that wearable devices should collect with explicit consent. The following categories of data are defined as HIPAA identifiers (comma delimited): Name, Address, All elements of dates related to an individual, Telephone numbers, Fax number, Email address, Social security number, Medical record number, Health plan beneficiary number, Account number, Certificate or license number, Vehicle identifiers and serial numbers, Device identifiers and serial numbers, Web URL, Internet protocol address, Finger or voice print, Photographic image, and Any other characteristic that could uniquely identify the individual.

### Appendix D.3. Metric Description — De-identifies Data When Sharing

Public datasets offer significant benefits by increasing the amount of available data, yet this often comes at the cost of individual privacy. Organizations frequently de-identify data—that is, remove sensitive personal information while retaining other useful details—to mitigate privacy risks^43^. De-identification performed at the time of data collection reduces the likelihood of exposing sensitive information, thereby enhancing user privacy protection. However, reidentification of anonymized data is emerging as a serious concern. A 2016 report documented a 320% increase in hacking attacks on healthcare providers, and the volume and detail of wearable data only exacerbate these vulnerabilities^44^. Conventional de-identification techniques such as k-anonymity^45,46^, l-diversity^39,47^, and t-closeness^39,45^ each present unique challenges in preventing re-identification, highlighting the need for continued innovation in privacy protection methods.

#### Recommended Performance

The vendor should de-identify the data before sharing with other entities.

By nature of the request, it would be impossible to determine what de-identification method is utilized by vendors. Any data that can be tied to health data should be de-identified before sharing.

### Appendix D.4. Metric Description — No Third Party Sharing

Most wearable manufacturers include clauses about sharing data with third-party vendors, who in turn may use the data for purposes outside the explicit consent provided by users. Numerous service policies evaluated in this review include general third-party data-sharing provisions from major vendors such as Apple and Google’s Fitbit^48–50^. In addition, few vendors explain which third-party services gain access to users’ personal data or why such access is granted; instead, they rely on broad, all-encompassing language to account for potential future actions^51^. Furthermore, many policies include provisions that allow sharing identifiable information when required by law, which could lead to harmful legal repercussions for users. Tech policy researchers in the United States have warned of the potential privacy invasions by period-tracking apps following the overturning of Roe v. Wade^52–55^. Overall, sharing data with third parties can pose significant risks to a user’s privacy.

#### Recommended Performance

The vendor should not share data with third parties.

### Appendix D.5. Metric Description — Transparency in Third Party Sharing

Vendors must fully inform users about all data sharing practices and the underlying intent. Ard (2013)^56^ argues that it is increasingly difficult to identify and regulate third parties because they are not obligated to adhere to the privacy agreements established between users and vendors. The study proposes that certain types of information should remain confidential regardless of who may acquire it. However, this approach necessitates that both users and regulators are aware of all third parties that receive user data, thereby requiring complete transparency and disclosure. Notably, a vendor that does not share data with any third party would automatically be considered fully compliant with these privacy standards.

#### Recommended Performance

The vendor should disclose all third parties that receive user data. Any language of data sharing with vague or unspecified third parties is considered not transparent.

In one study, human annotators determined the top 5 vague words in privacy policies to be “may”, “personal information”, “information”, “other”, and “some”^57^. Another study analyzed mobile health and fitness apps to find that privacy policies were frequently not applied to third-party links and services^58^. One researcher goes as far as to coin the term “Incognito Problem” to refer to users being provided little to no information on which third parties obtain user data^59^.

If a vendor does not share data with third parties, the vendor is awarded this metric automatically.

### Appendix D.6. Metric Description — Secure Wearable Connectivity

Wearable devices rely almost entirely on wireless communication—a technology that began gaining traction before the turn of the millennium^60^ and has since become the primary method for device-to-device connectivity^61,62^. A variety of wireless protocols, such as WiFi, ZigBee, Z-Wave, Sigfox, Neul, LoRaWAN, RFID, NFC, GSM/3G/4G, Bluetooth LE, GLoWPAN, HomePlug, Thread, DSRC, and WiMax, are used by IoT devices, each presenting different levels of security vulnerabilities^63,64^. Among these, Bluetooth LE is the most popular and practical communication protocol for the majority of wearable devices on the market^65,66^. However, vulnerabilities in these wireless connections or in pre-authorized smartphone applications can be exploited by malicious actors to access sensitive data^67–70^. Between Bluetooth LE and WiFi, Bluetooth LE is favored for its lower energy consumption and suitability for resource-limited devices, but it is vulnerable to man-in-the-middle attacks which can lead to significant security risks^68^. Common Bluetooth attacks include bluesnarfing, eavesdropping, and packet injection^68,69^. Since wearables often serve as entry points for hackers, compromising these devices can also expose smartphones or computers to broader security breaches^71^.

On the other hand, WiFi is generally considered more secure than Bluetooth despite both employing 128-bit encryption, because WiFi tends to employ certificate-based, server-backed authentication frameworks (WPA2 or WPA3 Enterprise)^67,72^. Although WiFi is not free from vulnerabilities—as seen in the 2016 botnet attacks that compromised millions of devices^73^—transmitting wearable data via WiFi is highly recommended. That said, the security of WiFi is highly dependent on the user-s network configuration. Users should ensure they connect through WPA-2 or WPA3 networks since WEP is known to have significant security weaknesses^74,75^. Users are also susceptible to remote attacks that would typically not be a risk for local connection protocols.

#### Recommended Performance

Wearable devices should be capable of connecting through WiFi.

Wearable devices use several methods to communicate, including ANT/ANT+, Near Field Communication (NFC), Bluetooth/Bluetooth Low Energy (BLE), Zigbee/Thread/Z-Wave, WiFi, and Cellular (3G/4G/5G). However, the security of networking protocols in wearable devices depends highly on both implementation and specific use case. Protocol-level defaults (i.e. design specifications) determine the base level for certain standards^76^, vendor implementation influence whether devices have encryption or robustly secure keys (e.g. assigning unique identifiers for advertising purposes–increasing risk of long-term tracking^77^), and client-sided factors such as a user connecting over an unencrypted network. Ranking network protocols is largely difficult due to the concerted nature of establishing a secure network (protocol, vendor, user/client) and a single point of failure in adherence of the three could expose vulnerabilities. However, the proposed ranking (from least to most secure) based on protocol characteristics, default settings, and other one-off considerations are as follows:

- **ANT/ANT+**: Not encrypted by default, no built-in authentication, AES-128 is optional and by way of single-channel communications^78,79^.
- **NFC**: Offers fast, secure pairing with man-in-the-middle resistance, but can be vulnerable to eavesdropping, data modification, and tracking^80^ especially if an attacker can get physically close–but also requires an attacker to get close^81^. No confidentially guarantees by default^82^
- **Bluetooth/BLE**: Although these protocols incorporate security measures, implementation is left to device manufacturers and there are known security issues^83^.
- **Zigbee/Thread/Z-Wave**: Security is dependent on the implementation by the manufacturer^84^.
- **WiFi**: Offers strong security, especially with WPA2 and WPA3, but because it is implemented locally it increases the potential for remote attacks^85^.
- **Cellular (3G/4G/5G)**: Managed by telecom companies that adhere to stringent security regulations, offering robust protection^86^.

### Appendix D.7. Metric Description — Secure API Access

Permissions to access data storages are typically provided through APIs or SDKs developed by device manufacturers. APIs serve as tools for applications to interact with each other, while SDKs package these API methods for similar functionality. These interfaces can be either public, allowing any internet-connected entity to access data, or private, restricting access to select entities within a controlled network. Many modern applications, including wearable platforms, depend on APIs for authentication and data access^87^. When a vendor offers a public API, users can download data stored on company servers from anywhere in the world^88^. However, before any data is accessed, authentication and authorization protocols are required to ensure that only authorized users gain entry^63^. Among the various protocols available, OAuth 2.0 has become the de facto gold standard for authorization due to its robust authentication and authorization properties, despite having its own security vulnerabilities^89^.

#### Recommended Performance

Vendors should utilize the OAuth 2.0 protocol (or more secure protocol, explained below) when exposing data through APIs.

Below is a list of common authentication and authorization protocols used for API data access ranked from least to most secure. This is followed by an explanation of which performance attributes are desirable.

- **Basic Authentication**: Sends Base64-encoded credentials with every request; vulnerable to man-in-the-middle attacks unless used over HTTPS^90^.
- **Digest Authentication**: Transmits a hashed password instead of plain text but still vulnerable to man-in-the-middle attacks and lacks password salting^91^.
- **API Key Authentication**: Assigns a unique key per user that is included in each API call; simple but risky if the key is exposed^92,93^.
- **OAuth 1.0**: Utilizes cryptographic signatures with a shared secret for authorization; more secure than earlier methods yet complex and largely superseded^94^.
- **OAuth 2.0**: The industry standard for authorization, offering tailored flows for various platforms and balancing strong security with developer simplicity^95–97^.
- **OpenID Connect (OIDC)**: Adds an identity verification layer on top of OAuth 2.0, enabling clients to confirm the end-user’s identity and access profile information^97^.
- **Mutual TLS (mTLS)**: Extends TLS by requiring both client and server authentication, providing the highest level of security despite higher computational overhead^98^.

Effective API access protocols must balance robust security with efficiency. OAuth 2.0 is widely adopted because they offer strong security without significantly impacting performance, making them suitable for modern, distributed applications. Thus, we recommend that vendors utilize the OAuth 2.0 protocol (or more secure protocols mentioned above such as OIDC or mTLS) when exposing data through APIs.

#### Privacy and Security Rating Citations

Sources for how each privacy and security rating was graded is provided in Table D.4. Most of the sources used to rate wearables were privacy policies, terms of use agreements, or manuals provided by each vendor. This is largely satisfactory due to the legally binding nature of these use agreements between users and vendors. While incidents have occurred of vendors violating these agreement policies, few if any third parties can accurately verify whether a vendor is violating their own terms of use agreements and is therefore a limitation of these ratings.

**Table D.4:**
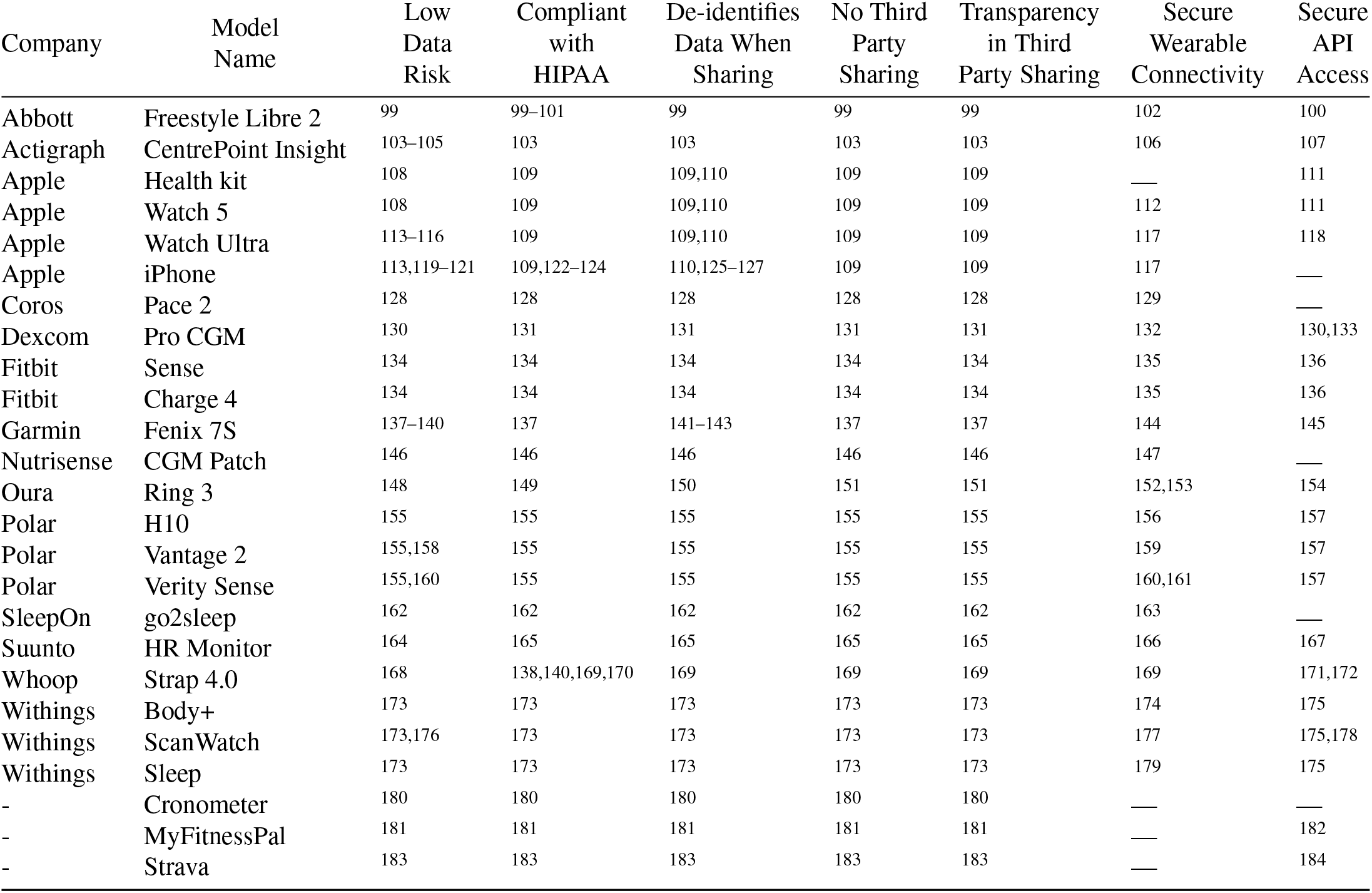
Citations for Table 3. Each source was accessed within the last year (2024 onwards) and scoured for language that provides evidence of meeting or failing the rating (as expressed under recommended performance in Appendix D). A dashed line indicates that result is not applicable or a source could not be provided.

